# The introduction of group antenatal care in northern Nigeria: an implementation research case study on improving quality and uptake

**DOI:** 10.1101/2025.09.07.25335045

**Authors:** Abdulhamid Abubakar, Grace Yisa, Adeola Seweje, Joseph Odu, Costa Atori, Uche Ralph Opara, Rondi Anderson

**Author notes:** (corresponding author) Rondi Anderson PhD, Project HOPE.

## Abstract

**Introduction:** With one of the highest rates of maternal mortality in the world, improving antenatal care for the poorest communities is a high priority in Nigeria. In 2022, Project HOPE partnered with the Nigerian Ministry of Health to implement a group antenatal care project in Niger State.

Government managers and Project HOPE aligned policy documents and budgets, and conducted a baseline assessment. Project staff then trained health care workers on group antenatal care. The project also connected women with a national health insurance scheme and strengthened monitoring and supervision systems.

**Methods:** Using quantitative implementation research, an instrumental case study was developed describing the design and implementation of the project. A final evaluation used both program and national health information system data to examine service utilization and health outcomes. Two tools were created for this purpose—a patient register and a monthly summary derived from that register.

**Results:** The project trained 300 health workers and established group antenatal care in 150 health facilities. Over 17,000 women participated in 1,054 antenatal care groups. Multivariate analysis found statistically significant increases in first and fourth antenatal visits and facility births following the intervention.

**Conclusion:** Despite challenges, this project demonstrated success in increasing service use. Influencing factors were the use of indigenous master trainers, engaging stakeholders, revitalizing facility monitoring and evaluation, and the inclusion of the group antenatal care model in national antenatal care guidelines. Noted challenges included inadequate skilled birth attendants and lack of antenatal care commodities.

- **What is already known on this topic** – *Maternal mortality is high in Nigeria. Antenatal care and facility birth are evidence-based practices for reducing maternal mortality. A large-scale group antenatal care project was implemented in 150 primary health centers in Niger State*.
- **What this study adds** – *We documented project implementation and analyzed changes in service use and health outcomes to share program challenges and impact. Despite funding limitations and a short implementation period, the project increased antenatal care attendance and facility births*.
- **How this study might affect research, practice or policy** – *The results show that group antenatal care implemented using a midwifery care model, and connected to state health insurance schemes and facility mentorship, can increase evidence-based practices that lower maternal mortality*.

## INTRODUCTION

Nigeria is responsible for 29% of global maternal mortality (1). With the third highest maternal mortality ratio of 993/100,000 and a population of over 232 million, maternal mortality is slow to decline (2,3). A major contributor to this is gaps in access to quality maternal health care, particularly for rural women. More than 15% of pregnant women in almost 2/3 of Nigeria’s states receive no ANC, higher than global averages (4). In 2016, WHO issued recommendations on ANC for a positive pregnancy experience (5). The recommendations prioritized “person-centered care” as essential for improved health and well-being. Communication and support at ANC contacts were highlighted as key to improving quality and utilization of healthcare services. Group antenatal care (G-ANC), a facility-based alternative model designed to address these priorities, was recommended as a health system strengthening intervention (6). This article is an implementation research case study of Nigeria’s introduction of G-ANC in 150 of Niger State’s 274 primary health care (PHC) facilities.

Niger State, in the North-Central region of Nigeria, has an estimated population of 5.9 million, of which approximately 1.2 million (22%) are women of reproductive age. While about 60.6% of Niger’s population is employed—90% in the informal sector—the State’s poverty incidence of 33.9% is lower than the North Central average of 59.7%. Niger has 25 local government areas (LGAs) with a total of 1,335 health facilities, of which two are tertiary, 21 are secondary, and the remaining are primary. Primary health facilities have a severe shortage of skilled human resources with associated gaps in functionality. Only 60% of women in Niger State receive any ANC care, despite the national adoption of the World Health Organization’s minimum of eight ANC contacts (4). Only 2% of women in Niger have full ANC coverage.

Niger is characterized by a widely dispersed population, with over 4,100 hard-to-reach settlements. According to the 2018 National Nutrition and Health Survey, Niger State has a Maternal Mortality Ratio (MMR) of 352 per 100,000 live births (7). Between 2011 and 2016, infant and under five mortality increased, and the number of births attended by skilled birth attendants (SBA) declined by one third. Notably malaria, a known contributor to maternal and neonatal mortality was found to have a prevalence of 27% in 2018 (8).

G-ANC is associated with improved care quality, higher ANC care seeking, and higher facility delivery rates (9). The G-ANC model is meant to empower women by enhancing peer-to-peer support and self-care. Groups share awareness raising, and pregnancy checkups, with some time for individual assessment with the health care provider. Leading into this project, Nigeria had conducted successful trials of G-ANC. The goal of this project was to upscale G-ANC availability in Niger State and thereby improve care quality and health seeking.

### Baseline Assessment

In preparation for this project a baseline facility readiness assessment was conducted. Interviews were held with eight policy makers followed by visits to 274 health facilities. Two hundred and sixty-four facility managers and health care providers, and 278 beneficiaries were interviewed. Almost all the targeted facilities were found to provide ANC, intrapartum care, and postnatal care (PNC), although hours were limited with only 67% providing 24/7 care. Ninety percent of health facilities had adequate space for ANC and 50% had adequate seating, hand wash basins with soap and water, and drinking water. Notably less than 10% had devices for measuring blood pressure. Only 6% of the health workers were skilled, with 94% being ancillary or community outreach staff. Seventy five percent of health workers had received reproductive, maternal, newborn, and child health training at some point in their career.

Focused antenatal care (F-ANC), or women meeting individually with providers, was the predominant ANC care model in the state. Forty eight percent of interviewed women had given birth to their most recent child at a PHC and 60% of pregnant women were planning a facility birth. Fifty five percent of pregnant women had at least one ANC visit with only 13% and 9% reaching the fourth and eighth visits. Seventy percent of pregnant women received at least one malaria prophylaxis treatment and 9% received insecticide treated bed nets. All facilities had data officers, and reporting was ongoing to various degrees. Additional findings included 1) inadequate skills 2) inconsistent supplies of drugs and other consumables and 3) fees for services affecting uptake including for facility birth.

## METHODS

### Study Design and Objectives

This study was part of a larger implementation project approved by the Ministry of Health and Family Welfare and funded by the Bill and Melinda Gates Foundation under project ID BGD10MWC. The primary objective was to understand the acceptability, practicality, and outcomes of G-ANC. An intrinsic case study format was used, employing mixed methods in order to synthesize and communicate project monitoring information (9).

### Patient Involvement

Women who had participated in the G-ANC intervention and their families were not involved in setting the research question or the outcome measures, but they were they intimately involved in design and implementation of the intervention. Women and their families were also central to dissemination of the baseline information, which helped to motivate community involvement during and beyond the study.

### Data Sources and Collection

Data collection and analysis were conducted by the project monitoring and evaluation team. Two tools were developed for this purpose: a register for G-ANC clients and a monthly summary derived from that register. The register tracked a range of service delivery and health outcome indicators, including training, number of cohorts, community outreach activities, first and fourth antenatal care visits, total and live births, malaria positivity among febrile covid patients, postpartum contraceptive use, and postnatal clinical visits for both mothers and newborns.

Quantitative data on the implementation of the program, and changes in service utilization after the initiation, were collected to understand the practicality and success of implementation. Data were collected from all 150 project facilities as part of program reporting, and analysis from existing government databases complemented project data. Data was collected on all women who attended maternity services at the facilities providing G-ANC. Quantitative data were sourced from the national health information system (DHIS2). Selected data was reported descriptively including G-ANC cohorts, and referrals from community health workers.

### Quantitative Analysis

To evaluate the impact of the G-ANC intervention, a multivariate interrupted time series analysis using linear regression was conducted using R version 4.5.0 (11). The model included eight outcome variables: ANC first visit, ANC fourth visit, number of deliveries, number of live births, proportion of malaria-positive cases among febrile patients, postpartum contraceptive use, postnatal clinical visits for mothers, and postnatal clinical visits for newborns.

### Project Description

#### Design

As part of a national initiative in May of 2022, Project HOPE with the Ministry of Health commenced the G-ANC project in Niger State. Inception and co-creation meetings with key stakeholders were held. An initial desk review of successful G-ANC practices was completed, and based on that, the national and state health policy, annual operation plan, and budget were revised to include a G-ANC model of care. To evaluate the project, monitoring and supervision systems were strengthened including a strong focus on quality data collection and reporting. For implementation of the project and accurate monitoring, key tools were developed, adapted, and contextualized. The tools included an orientation package for local health authorities, a harmonized mentoring checklist, job aids for implementation of G-ANC, and a scheduling tool for planning G-ANC groups as prior to this initiative ANC was only by drop in. Data collection registers and summaries were used.

To maximize impact, collaborations were formed with government and partner initiatives that complemented G-ANC. The collaborations included the Malaria Consortium, the World Bank Accelerated Nutrition Program, Plan International, state programs for financing health facility equipment, state health insurance schemes, the Department of Health Planning Research and Statistics, and community outreach workers. These complementary programs distributed supplies for malaria prevention, ensured the sustainability of critical equipment for ANC, increased enrollment in health insurance, and supported community outreach. With these organizations, Project HOPE advocated to strengthen facilities with weaker services. This included bringing community-based services to facilities to ensure a larger reach, and training partners on quantification for the needed commodities. The State Health Education unit and media were also involved in key messaging for visibility and proper utilization through airing of jingles in indigenous languages.

#### Implementation

In September of 2022, training of health care workers (HCWs) on the provision of G-ANC was initiated followed by the roll out of the new G-ANC model. As the initiation of G-ANC started immediately after training HCWs it was staggered from September and November 2022. Enabling environments for care provision were strengthened by networking and synergizing with partners also focused on the health of pregnant women. G-ANC was implemented for 6 months from September 2022 to February 2023 at which time it was abruptly stopped due to funding issues. Over the 6 months of implementation, 301 health workers were trained and G-ANC was initiated in 150 health facilities. Enabling environments for care provision were strengthened through mentoring and by networking with national schemes for enrolling women in health insurance and supporting provision of malaria medicines. To evaluate the project, monitoring and supervision systems were strengthened.

#### Training, support, mentoring, and supervision

Several types of training at both state and local levels were initiated. They included training for policy makers, data collectors, community workers, health care providers, supportive supervisors, mentors, and managers. In addition to training, the project provided technical support and facilitated skills transfer to local health authorities. Ongoing quality improvement systems, mentoring, and supportive supervision were strengthened. Trainings included:

- Data management Project HOPE trained all 274 government designated monitoring and evaluation focal persons on DHIS2 and data management. Data management training was also conducted for 1,051 HCWs and managers.
- Community outreach Four hundred and seven community outreach workers, promotors and supporters (30 per LGA), were trained on demand creation and referral to G-ANC implementing facilities. The training utilized a variety of modalities including presentations, role plays, group work, and plenary sessions. In the end, the participants expressed confidence. In addition, advocacy visits and dialogues were conducted for community gatekeepers, including district heads, and women and youth leaders.
- G-ANC provision Training for provision of G-ANC included training and certifying 40 local master trainers in two batches, each for a duration of five days. These trainings built capacity on training trainers who then trained HCWs to implement the new model of ANC care. The newly trained trainers cascaded the training to 301 HCWs including 13 community health officers, 249 community health extension workers, and 31 nurse-midwives, two per health care facility. In addition, 25 MCH coordinators, and 15 PHC directors received the training. In the end 151 primary care facilities had trained health care workers. G-ANC facilitators were trained on the G-ANC methodology, and to encourage pregnant women to attend ANC visits, delivery in a facility, and enroll in G-ANC. The training and following implementation of G-ANC was conducted in phases. In the first month of implementation 112 HCW from 5 LGAs were trained, this resulted in 34 PHCs establishing G-ANC. In the second month, an additional 109 HCW from 57 facilities were trained and initiated implementation, in the third month health workers from 46 facilities were trained, and in the fourth and fifth month 13 more health facilities received the needed training. Following the trainings the master trainers made visits to the health facilities on ANC clinic days to provide hands on support enrolling women into G-ANC and conducting the sessions.
- Mentors and supportive supervisors To ensure enabling environments and support problem solving, 15 PHC directors and five state supportive supervisors were trained on G-ANC supervision. Additionally, 40 government maternal and child health coordinators and monitoring officers were selected and trained as mentors. They were existing local government leaders who took on an expanded mentoring role. To train the mentors and supervisors, a Project HOPE program manager and senior technical officer led a one-day meeting with key government stakeholders. Mentors and supervisors reviewed a visit checklist and capacity-strengthening indicators. All facilities were visited by mentors at least monthly and mentoring checklists were filled out at every visit to guide as a job aid and provide feedback to the program.

#### Supporting logistics and supplies

Facility officers in charge were trained and given technical support on accessing the Basic Health Care Provision Fund under Nigeria’s National Health Act, which has been designed to improve primary health care. The scheme includes resources for procurement of essential lifesaving commodities including blood pressure devices. Through this initiative, it was hoped that all health facilities would sustainably have at least one blood pressure cuff, though the baseline assessment results demonstrated otherwise. Improving supply chain management for commodities and equipment was thus prioritized through training and technical support for proper quantification, procurement, and distribution of commodities. In addition, some blood pressure cuffs were donated by the project to fill gap until all government provided supplies could be distributed. The project also copied over 8,000 government data collection tools, teaching materials, and service tracking forms for pregnant women.

#### Implementation challenges

Most challenges involved insecurity. Niger State has been plagued with kidnappings, banditry, cattle rustling and armed robbery particularly in certain locations. Some HCWs needed to join training in safer areas to protect the intervention teams, and a few HCWs and facilities could not participate because of the security concerns. The gaps in resources identified in the baseline assessment affected the project implementation. They included inadequate human resources, particularly midwives and nurses, and availability of essential equipment and supplies. Initiatives to leverage state programs for facility procurement of much needed commodities met barriers, and in the duration of the program only saw small success. Women were not aware of the much-needed government health coverage, and adequate support for enrolling all communities was not present. Because of gaps in resources and inadequate coverage, many very poor women need to pay out of pocket for basic health care, perpetuating hesitance to seek lifesaving interventions. In addition, outreach workers were not compensated on schedule, causing demotivation and interfering with aspects of program implementation.

## RESULTS

Ninety-one health facilities started implementing G-ANC in the first two months of the project and 137 in the first three. During the period of implementation, 33,883 pregnant women attended a first ANC visit (ANC1) of which 17,706 (52%) were enrolled in G-ANC. Only in the final months did the percentage of women receiving G-ANC exceed 50% of all women attending ANC. Conventional ANC was ongoing through implementation. The addition of mentors in the second and third months improved G-ANC attendance as it built confidence among the HCWs and managers. By the end of the project, a total of 17,706 women received G-ANC and over 1,000 G-ANC meetings occurred monthly. Meetings provided reliable peer-to-peer support, allowing for increased self-efficacy among pregnant women.

Out of the 301 HCWs trained to lead the G-ANC, 249 were community health extension workers; only 31 were nurses or midwives. Of the 407 government community outreach workers who were trained to increase awareness and refer pregnant women to the new G-ANC, only 72% ever referred a pregnant woman to G-ANC and by the end of the project only 16% were active. Overall, the outreach workers only contributed 6% of the total G-ANC attendance. Low and irregular payments to outreach workers interfered with their optimal participation.

The interrupted time series analysis model included three predictors: time (to estimate the pre-intervention trend), level (to estimate the immediate change after the intervention), and slope (to estimate the change in tren d following the intervention). To account for the staggered roll out across health facilities, the level and slope change variables were aligned with each facility’s specific implementation date. This approach allowed for estimation of the average intervention effect across sites while maintaining accuracy regarding the timing of implementation at each facility.

Figure 1 presents trends in each of the eight outcome variables over time, with a shaded region indicating the staggered start of the intervention across participating facilities (September– November 2022).

**Figure 1.**
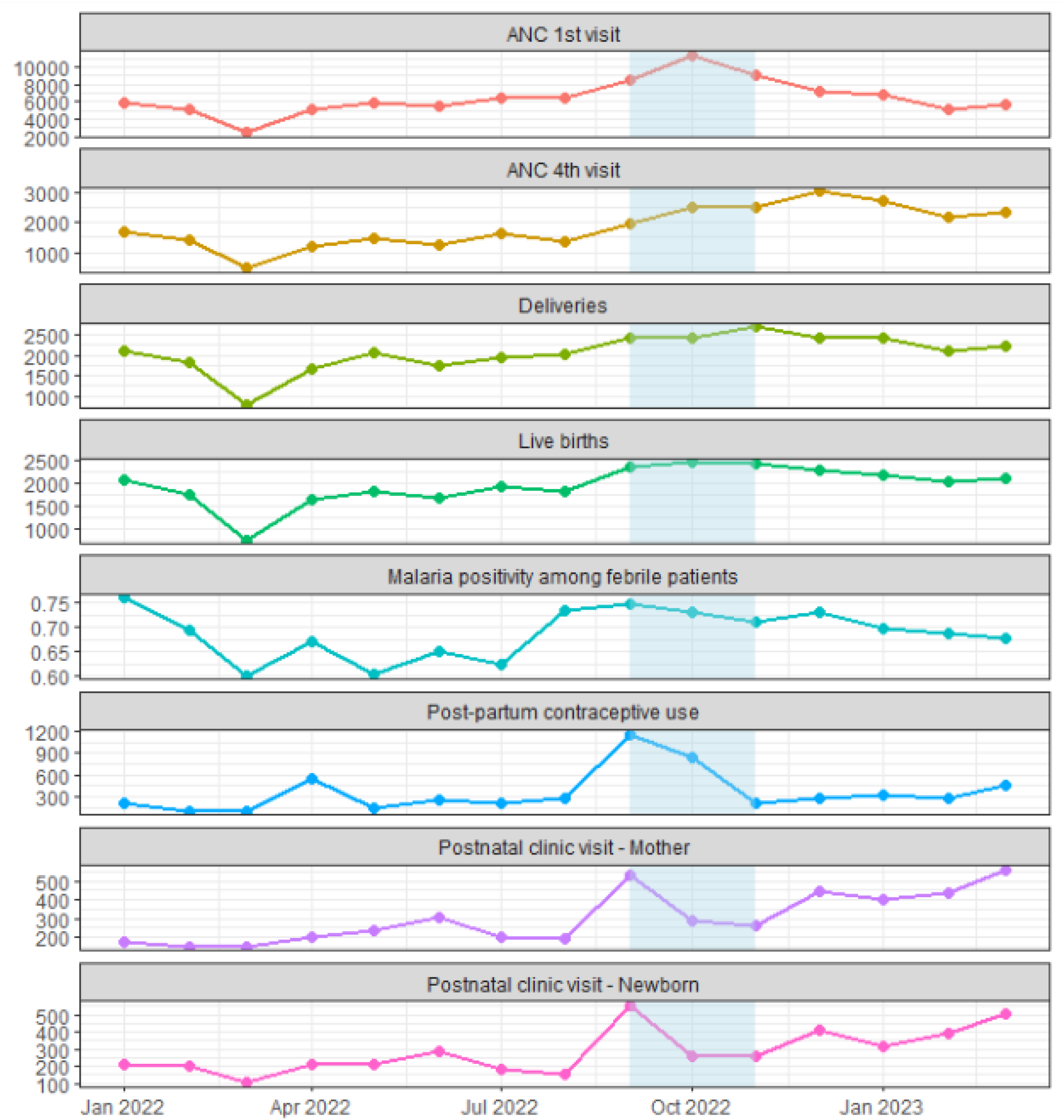
Trends in outcome indicators over time Note: Shaded region indicates period of intervention implementation.

A multivariate test indicated statistically significant effects for all three interrupted time series model predictors (Table 1). There was a significant effect of time (p < 0.001), indicating that trends were already changing prior to the intervention. In addition, both level (p = 0.027) and slope (p < 0.001) changes following the intervention were significant, suggesting that the introduction of G-ANC had both an immediate and increasing impact on key outcomes.

**Table 1.** Multivariate interrupted time series model results.

| Term | DF | Pillai Test Statistic | Approx. F | Num. DF | Den. DF | p-value |
| --- | --- | --- | --- | --- | --- | --- |
| **time** | **1** | **0.0417** | **4.480** | **8** | **824** | **<0.001\*\*\*** |
| **level** | **1** | **0.0207** | **2.181** | **8** | **824** | **0.027\*** |
| **slope** | **1** | **0.0552** | **6.017** | **8** | **824** | **<0.001\*\*\*** |

Follow-up univariate tests identified significant changes in four individual indicators: ANC first visit, ANC fourth visit, deliveries, and live births (Table 2). For ANC first visits, there was a statistically significant increasing trend before the intervention (p < 0.001), an immediate increase in level following implementation (p = 0.013), and a significant decline in slope (p < 0.001), indicating a slowing in the rate of increase post-intervention. ANC fourth visits also showed a significant level increase following the intervention (p = 0.004). Both deliveries and live births experienced significant level increases after the intervention (p = 0.028 and p = 0.033, respectively). There was also a significant time trend in postnatal clinical visits for mothers (p = 0.027) suggesting an increasing trend that started before the intervention.

**Table 2.** Univariate interrupted time series model results for each outcome.

| Outcome | Term | Estimate | SE | p-value |
| --- | --- | --- | --- | --- |
| ANC 1<sup>st</sup> visit | **time** | **3.06** | **0.805** | **<0.001\*\*\*** |
|  | **level** | **16.436** | **6.634** | **0.013\*\*** |
|  | **slope** | **-9.147** | **1.549** | **<0.001\*\*\*** |
| ANC 4<sup>th</sup> visit | time | 0.043 | 0.409 | 0.917 |
|  | **level** | **9.777** | **3.371** | **0.004\*\*** |
|  | slope | -0.517 | 0.787 | 0.512 |
| Deliveries | time | -0.309 | 0.396 | 0.436 |
|  | **level** | **7.209** | **3.265** | **0.028\*** |
|  | slope | -0.36 | 0.762 | 0.637 |
| Live births | time | -0.254 | 0.366 | 0.488 |
|  | **level** | **6.447** | **3.018** | **0.033\*** |
|  | slope | -0.36 | 0.704 | 0.609 |
| Malaria positivity among febrile patients | time | -0.009 | 0.006 | 0.123 |
|  | level | 0.092 | 0.049 | 0.061 |
|  | slope | 0.004 | 0.011 | 0.736 |
| Post-partum contraceptive use | time | 0.882 | 0.598 | 0.141 |
|  | level | -1.119 | 4.925 | 0.820 |
|  | slope | -1.194 | 1.15 | 0.299 |
| Postnatal clinical visit - Mother | **time** | **0.305** | **0.137** | **0.027\*** |
|  | level | -1.173 | 1.131 | 0.300 |
|  | slope | -0.063 | 0.264 | 0.812 |
| Postnatal clinical visit - Newborn | time | 0.219 | 0.133 | 0.101 |
|  | level | -0.882 | 1.099 | 0.423 |
|  | slope | -0.119 | 0.257 | 0.642 |

Other outcome variables, including malaria positivity among febrile patients, postpartum contraceptive use, and postnatal clinical visits for newborns—did not show statistically significant changes during the study period.

## DISCUSSION

This project highlights the significant gaps in primary health care availability, quality, and service utilization that are common in low-resource settings, and sheds light on effective quality improvements. Despite challenges, this relatively brief G-ANC project made a difference. First and fourth ANC visit attendance increased, as did facility births and post-natal visits. While increases in certain measures were noted prior to roll out in Niger State, this was likely due to national level activities, such as the adoption of WHO guidelines, being initiated before state level roll out. Limitations to the project included local insecurity, inadequate skilled health workers, significant supply gaps, ongoing fees for services, and abrupt closure due to the stoppage of funding. The impact was in part a result of leveraging all stakeholders and related programs and building a strong network, but also demonstrated that G-ANC can improve quality of care and empower health workers and women, leading to improved care seeking (12,13,14).

The baseline assessment found that although the health system was in place, and certain aspects of it were functional, critical gaps in life-saving interventions, including essential equipment to prevent maternal death, and 24/7 care availability, were ubiquitous. Research exposes similar findings in many low-resource settings. Using demographic and health surveys from 10 low- and middle-income countries, Benova et al. (2018) found that in the poorest settings only 10-15% of health facilities provide the basic components of ANC (15). However, in the Benova et al. article, Nigeria ranked higher than other countries with rates of functional ANC as high as 50%. Globally, blood pressure monitoring is the most common component of ANC. In our baseline study, availability of blood pressure equipment had significant gaps.

Synergizing with many partners strengthened the project. This aspect of implementation is often overlooked for a number of reasons (16). Challenges that interfere with synergizing include fragmentation within weak health systems and competition between partners. Although all these concerns existed in this context, the project maximised the benefits of collaboration and benefitted from partner activities.

Mentorship also strengthened the project by supplementing training, which often falls short of expectations (12). Implementation commenced before mentoring and results improved with mentoring due to improved provider confidence. Mentors are acknowledged in the literature as effective for improving quality implementation and facilitating improvements in clinical care in low- and middle-income countries (13). Using existing government staff in the role of mentors has a precedent in literature and has been found to contribute to quality programming (14). Although there are advantages to using mentors external to the system, who are not burdened by other duties, and may be less influenced by political influences, using those within the system is more likely to support sustainability. The literature finds that mentoring can comprise both clinical and facility-wide interventions aimed to capacitate and create enabling environments for quality care (13). Mentoring enlists advocacy, modelling, and problem solving to achieve quality improvements.

While this project both directly provided blood pressure cuffs to facilities, it also worked on strengthening the national supply chain. Projects often struggle with the dilemma, highlighted in the health system strengthening literature, of donating commodities separately from government’s supply chain, as donations in general are not sustainable and may even demotivate system strengthening (17). This project was innovative in that it combined both approaches.

In the poorest countries there is a precedent to use community health workers who have at least 3 months training to provide ANC and skilled birth attendance in contexts without the global standard of provider. Their contribution to reducing morbidity and mortality, however, is unclear (18). What is clear is that engaging community health workers for outreach and referral does increase the rates of skilled birth attendance (19). While this analysis did not disaggregate outcomes by provider type, nor did it look at maternal and neonatal mortality, it did find a decrease in malaria and an uptake in family planning methods. Both of these are associated positively with maternal and newborn health outcomes. How to best address the needs of girls and women in contexts without standard educated health care providers such as midwives, is a real concern. More research is needed to understand the pros and cons of using lesser trained community health workers in this capacity, but this article suggests there are benefits.

The global recommendation for ANC and skilled birth attendants is midwife or equivalent who has completed a pre-service education meeting the standard defined by the International Confederation of Midwives (20). Nigeria has a national task shifting and task sharing policy focused on improving the quality of primary health care (21). The aim of the policy is to decentralize health care, as currently HCW availability is uneven, with notable gaps in rural areas. This enables community health workers to provide ANC services, and community outreach workers to connect pregnant women in communities with services. While there were no concerns noted with community health workers providing ANC, government community outreach workers linked few women to services largely because of constraints around their salaries. Notably, the literature finds mixed results for community outreach interventions (22,23,24). Community health workers have been shown to increase both facility birth and skilled birth attendant rates, but these results are more likely to be found where they are well supported by the system, which was not the case in this project.

The strong emphasis on data collection, monitoring and evaluation was an important aspect of the project. The project leveraged existing government employees to strengthen data management through a DHIS2 tracker who were active, collaborative partners. All pregnant women were able to be followed from registration through the post-partum period. While project monitoring is essential to understanding its impact, it is often limited by weak host country data collection systems. This project stressed the importance of continuous monitoring to ensure quality and worked closely within the government systems to ensure sustainability. This focus is recognized in the literature as a component of ensuring quality implementation (25,26).

This project had an unanticipated finding with regard to scheduling tools. Without a scheduling tool it was difficult to facilitate the G-ANC. The need to introduce a scheduling tool to improve quality or expand on ANC is found in the literature (27). No scheduling tool is often an unanticipated barrier to making changes in the more common system of drop-in care in low-resource settings, often associated with long waits. Although scheduling does facilitate more efficient care, particularly for the beneficiary, functionalizing scheduling tools in low-resource settings has complexities including that many women coming for care may not have smart phones so the system needs to be hybrid, and also that if the larger health system is not using a scheduling system, the providers themselves are burdened with negotiating the logistics.

## Conclusions

In spite of numerous limitations, including an only 6-month implementation period, this project saw care seeking and certain health outcomes improve. Through working within the government system and leveraging synergies, the project established a feasible and sustainable path to quality ANC. Strengthening the health system in the areas of data, service delivery, supplies and equipment, and social insurance, made it possible to successfully implement a G-ANC service delivery model.

## Supporting information

RECORD Checklist

## Data Availability

All data produced in the present study are available upon reasonable request to the authors

